# Unraveling dynamic factors in Dialectic Behavioural Treatment for Adolescents (DBT-A): a study protocol

**DOI:** 10.64898/2025.12.18.25342603

**Authors:** Anneke de Weerd, Anne Krabbendam, Agaath Koudstaal, Joep Sins, Jacquelijne Schraven, Robert Vermeiren, Jantine Roeleveld, Elisabeth Koopman-Verhoeff, Laura Nooteboom

## Abstract

Dialectical Behavioural Therapy for Adolescents (DBT-A) is an effective treatment for adolescents exhibiting features of borderline personality disorder (BPD). However, some do not benefit, potentially leading to major negative outcomes later in life. Previous quantitative studies have highlighted the importance of dynamic (changeable) factors in determining treatment success, including the therapeutic alliance, emotional dynamics between therapist and adolescent, and the motivation of those involved. Nevertheless, the specific contribution of these factors to the therapeutic process and perceived treatment success remains unclear. This study protocol outlines a multicenter, qualitative longitudinal study designed to explore how dynamic factors in DBT-A are experienced by adolescents, parents, and therapists at various stages of therapy. The study will track treatment journeys (trajectories) of fifteen youth during DBT-A therapy across four child and adolescent psychiatry institutions in the Netherlands. The triad of adolescent, parent(s), and therapist will be interviewed separately at four time points during treatment: at the start and after three, six, and nine months (resulting in 180 interviews in total). The semi-structured interviews, based on a theoretical framework, will be analyzed using reflexive thematic analysis, employing a combination of deductive framework analysis and inductive open coding methods. By bringing together the triad of perspectives, and monitoring how these perspectives evolve over time, this study will yield valuable, practice-based insights into how dynamic factors unfold during DBT-A. Insights from this study will offer therapists specific, actionable guidance for modifying dynamic factors to better address the unique needs of adolescents with BPD features.

## Introduction

Dialectic Behavioural Treatment for Adolescents (DBT-A) is well-established as an effective intervention for reducing self-harm, suicidal ideation and suicidal behaviour in adolescents with features of borderline personality disorder (BPD) [1, 2]. When followed for a longer period, DBT-A is also effective in reducing other BPD symptoms [3]. However, these positive outcomes are typically demonstrated at a group level, and a proportion of adolescents at risk for BPD do not respond or show insufficient improvement to this evidence-based treatment. This is important, as BPD is known as a long-term psychiatric condition with lasting negative effects on both the individual and their surroundings [4]. Additionally, this is a group known for high healthcare costs and long-term care needs within a system that is already under pressure [5]. Accordingly, early and effective intervention is essential.

Both empirical studies and clinical practice highlight that the effectiveness of BPD is strongly influenced by dynamic (i.e. changeable) factors, such as the quality of the therapeutic alliance, the capacity to experience and address ruptures in the therapeutic relationship and the alignment of mutual expectations [6–9]. Despite their importance, there is a notable gap in knowledge regarding how these dynamic factors are experienced and perceived by adolescents, their parents/caregivers (hereafter referred to as parents) and therapists over the course of treatment, and how these factors develop over time during treatment. Therefore, the present qualitative, longitudinal and multi-perspective study aims to investigate how dynamic factors in DBT-A are experienced by adolescents, parents, and therapists at various stages of therapy.

### BPD features in adolescents

Borderline Personality Disorder (BPD) is characterised by unstable interpersonal relationships, a disturbed self-image, impulsivity, and severe mood swings [10]. Although there has historically been some clinical reluctance to diagnose BPD during adolescence, an increasing body of international research supports the importance of early identification and intervention [11, 12]. Adolescents presenting BPD features often experience significant impairments across multiple domains, including severe emotional dysregulation, difficulties in forming social relationships and school problems [13]. Moreover, they frequently engage in suicidal behaviours, non-suicidal self-injury, low self-esteem, and behavioural problems including running away and substance misuse [4, 14, 15].

Several psychosocial factors contribute to the development of BPD symptoms in adolescents, including persistent family conflicts, feelings of emotional disconnection, and the internalized belief of being a burden to others, all of which serve to reinforce maladaptive patterns [16]. Consequently, many adolescents with BPD struggle to develop stable peer relationships and successfully complete their education, leading to compromised long-term social and occupational outcomes [13]. If left untreated, BPD in adolescence can result in poor socio-economic outcomes and further dependence on mental health systems [5].

### Dialectical Behavioural Therapy for Adolescents (DBT-A)

BPD symptoms can be significantly reduced through Dialectical Behavioural Therapy for Adolescents (DBT-A) [3, 17]. DBT-A is typically provided over a six to twelve month period, within specialized child and adolescent psychiatric settings. Treatment consists of multiple integrated components, including skill-training groups, individual therapy, telephonic consultation and system (family) therapy [2, 18]. Therapists are participating in a consultation team.

DBT-A aims to improve adolescents’ abilities in distress tolerance, mindfulness, and emotion regulation, foster a stronger sense of connection with others, and reduce experiences of interpersonal rejection and conflict. Involving parents, and when appropriate the broader social network is considered a fundamental element of DBT-A, distinguishing it from the adult version of DBT where such involvement is not standard [18]. This component is particularly important because many adolescents with BPD features report pervasive feelings of disconnection and burdensomeness toward their parents or caregivers [19]. At the same time, caregivers often struggle to provide emotional support in ways that foster the adolescent’s sense of belonging and self-worth [16], while also experiencing significant distress due to caregiving challenges associated with their child’s borderline symptoms [20].

Although DBT-A has demonstrated overall effectiveness for adolescents with BPD features, treatment, outcomes can vary considerably [3, 17]. In some cases, poorer treatment outcomes— such as limited symptom improvement, ongoing self-harm, emotional dysregulation, or lack of engagement—contribute to premature dropout from treatment [3, 21].

Existing research indicates that treatment success in BPD is influenced by both static (unchangeable) and dynamic (changeable) factors [7, 22, 23]. Static factors such as biological sex and intelligence quotient may help identify which individuals are more likely to benefit from treatment, but cannot be directly influenced by therapeutic interventions. In contrast, dynamic factors, defined by their potential for change, are often shaped through the interactions between therapist, adolescent and their surroundings throughout the course of treatment. These dynamic factors are particularly relevant for understanding different outcomes and for optimizing therapeutic strategies in DBT-A.

### Previous research on dynamic factors

Various dynamic factors have been identified as contributing to treatment success for adolescents with BPD features. These include the quality of the therapeutic alliance [8, 9], the emotional responses that adolescents and therapists hold toward each other [5, 24], and the ability to recognize, experience and repair ruptures in the therapeutic relationship [25]. Additional factors are the adolescent’s expectations regarding treatment outcomes [26], their sense of connection to others [18] and to themselves [24], the development of emotion regulation skills [8] and the opportunity for personal disclosure within therapy [27].

Beyond the context of BPD specifically, other dynamic factors have also been shown to influence therapeutic outcomes. These factors include therapist responsiveness, the attitudes of those involved toward treatment and problematic behaviours, the therapist’s communication style, and the patient’s motivation and commitment to psychotherapy [28–30]. Despite growing recognition of their importance, little is known about how these dynamic factors evolve throughout the course of treatment and how such changes contribute to positive or negative outcomes. Understanding temporal changes, enables clinicians to identify critical intervention points and tailor therapy to individual patient needs, thereby potentially optimizing treatment outcomes. Remarkably, only one study has examined helpful and unhelpful therapist behaviours in DBT-A from the perspective of former adolescent patients [31]. This study highlighted, among other elements, the therapist’s ability to convey belief in adolescent’s capacity to change destructive behaviour as a crucial factor. However, the retrospective nature of this study limited the ability to capture the evolving nature of therapeutic processes over time.

Additionally, parental involvement, a core component of DBT-A, has also been shown to positively affect treatment outcomes for both adolescents and their parents [31–33]. Nevertheless, how parental involvement interacts with dynamic factors in treatment, and what parents themselves need during treatment to support perceived treatment success for both themselves and their child, remains unexplored.

### The present study

In sum, dynamic factors – such as the therapeutic alliance and the adolescent’s motivation – are known contributors to DBT-A effectiveness. However, there is limited understanding on how these factors emerge and develop during treatment, and ultimately impact perceived treatment success. To address this gap, the present qualitative, longitudinal study aims to explore how dynamic factors in DBT-A are experienced by the perspectives of adolescents, their parents, and therapists over the course of nine months following the start of DBT-A treatment. In addition, the study will examine the role of parents during the treatment process. By capturing the experiences of adolescents, parents and therapists across multiple treatment trajectories, and by identifying patterns of change and contextual factors that shape them, this study aims to deepen our understanding of how dynamic factors contribute to perceived positive and/or negative treatment outcomes in DBT-A.

## 2. Methods

### Study design

This study adopts a prospective, longitudinal multiple-case design to explore how experiences and perceptions of dynamic factors in DBT-A evolve over time across multiple paths (trajectories) including adolescents, their parent(s) and their individual therapist from multiple institutions. This design enables in-depth examination and comparison of individual cases, allowing for the identification of distinct patterns of change over time [34]. Furthermore, this study aims to explain how and why these changes occur, with the broader objective of understanding the mechanisms that contribute to perceived treatment success or treatment failure. Four centres participate in this multi-centre study: all four specialized child and adolescent psychiatric institutions in the Netherlands that provide DBT-A treatment: Altrecht, Levvel, LUMC Curium and Youz (Parnassia Group). These institutions utilize the Dutch manual *Surfen op Emoties* [18] as the standard skills training book for DBT-A. Each site is actively involved through participation of a DBT-A therapist in the project group, contributing to both the development and implementation of the study. Each institution will contribute approximately three to four cases, ensuring a diverse sample across treatment settings.

Each case will include three key informants: the adolescents receiving DBT-A, their parent(s), and their individual therapist. Participants will take part in individual semi-structured interviews at four preplanned time points:

- T0: Start of treatment (immediately after commitment phase)
- T1: Three months into treatment (midway point)
- T2: Six months into treatment (completing treatment or deciding to extend)
- T3: Nine months (finished or halfway through therapy extension)

These time points have been strategically selected to capture critical moments of change, enabling analysis of both the processes and perceived impact of dynamic factors within DBT-A over time (see Figure 1). In addition to the interviews, participants will complete a brief questionnaire at each time point. These data will be used to supplement and contextualize the qualitative findings, offering a more comprehensive understanding of perceived treatment success.

**Figure 1.**
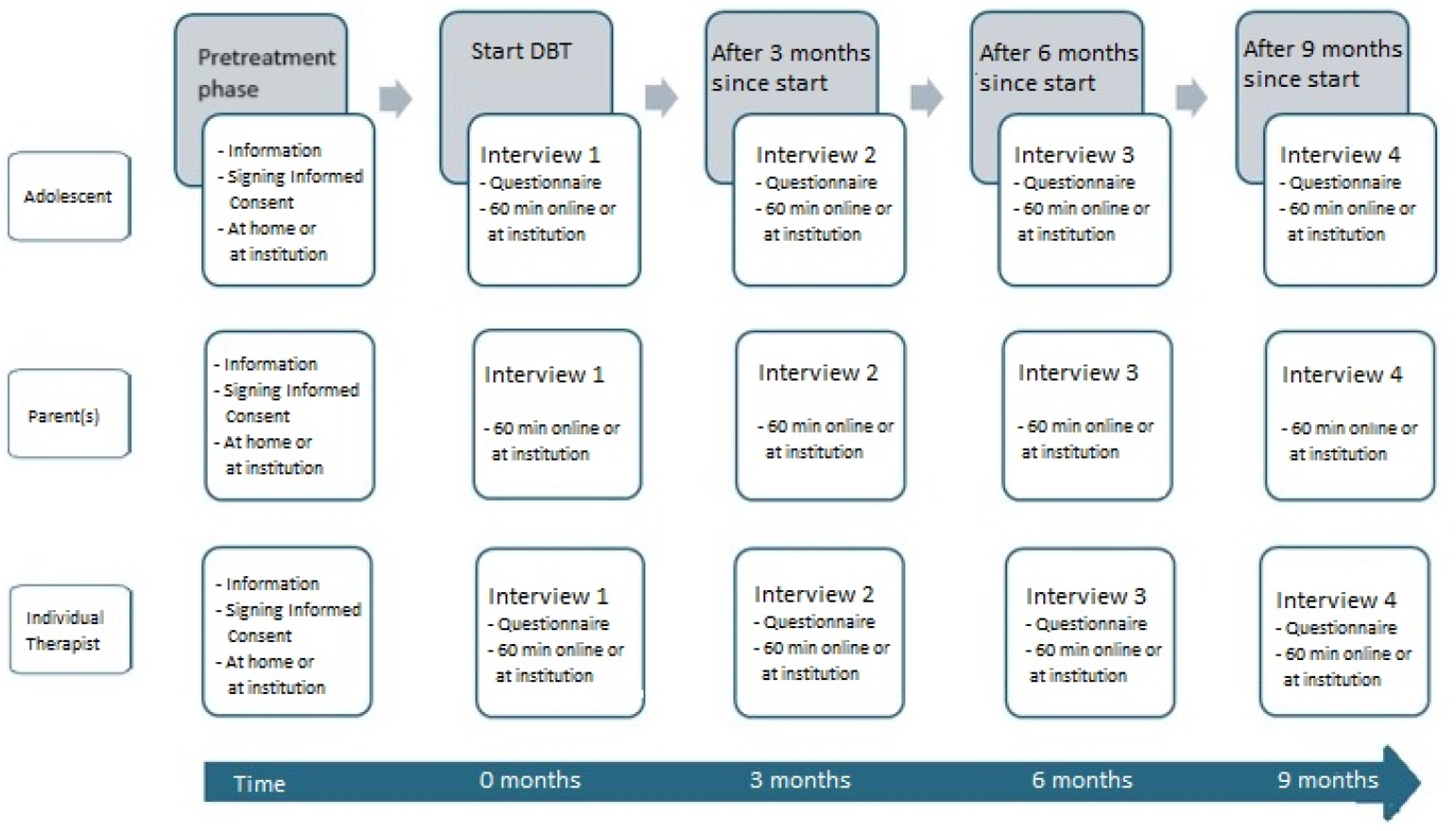
Visual overview of the study procedure

### Participants

The study will include approximately 15 cases, each consisting of one adolescent receiving DBT-A, their parent(s) or primary caregiver(s), and their individual therapist. Participants are recruited from four specialized, outpatient and day treatment programs offering DBT-A. To be eligible, adolescents must meet the following criteria: (1) Be between 12 and 23 years old at the start of treatment; (2) Be enrolled in a DBT-A program (either outpatient or day treatment); (3) Meet at least three diagnostic criteria for BPD according to the DSM-V (9), measured by the Mclean Screening Instrument for Borderline Personality Disorder (MSI-BPD) [35], meaning that both adolescents formally diagnosed with BPD and those presenting with BPD features are eligible. DBT-A is delivered to adolescents with persistent features of BPD, as established by routine clinical assessment at each participating institution prior to treatment initiation. For this study, MSI-BPD will be administered to record the specific DSM-5 criteria met and to confirm that at least three criteria are present. As eligibility is already verified through standard clinical procedures, completion of the MSI-BPD is the only additional requirement for study inclusion. Adolescents with comorbid ASD are not excluded, allowing for examination of their potentially distinct response patterns to DBT-A.

All parents or caregivers actively involved in the adolescent upbringing – whether biological, step, adoptive or foster – are eligible to participate and will hereafter be referred to as ‘parents’. Where possible, both parents will be interviewed together in a dyadic interview. If this is not feasible, the participation of one parent will be sufficient.

Therapists must be trained in DBT-A and employed as individual therapists within the participating institutions; in case of therapist unavailability (e.g., illness, staff turnover or parental leave), a replacement therapist will be invited to participate, and if they consent, the study will continue with their involvement. These changes during the course of treatment will also be monitored as part of the research data.

Inclusion of a case requires participation from the adolescent, parent(s) and the individual therapist. If any of these participants declines to participate, the case will be excluded from the study. Non-participation will have no consequences for the adolescent’s treatment, and participation in the study is entirely voluntary. In cases where the adolescent discontinues treatment prematurely, the adolescent, their parent(s) and therapist will be invited to participated in one final interview to reflect on and evaluate treatment experience. The interview schedule will be adapted accordingly, with the final interview conducted after treatment termination. Depending on individual circumstances, the interview will occur either as soon as possible or after a designated interval post-treatment.

The final number of participants will be based on the principles of thematic saturation [36]. It is expected to distill the core themes from this number of interviews (n=180) to answer the research questions and thematic saturation. Our purposive sampling approach is designed to ensure a representative and clinically relevant sample [37]. Transferability will be increased by conducting research in four institutions and paying attention to the diversity of the participants [38], reflecting gender, age and clinical presentation seen in practice [37]. Whether sufficient information has been found about the core themes will be determined after no new information occurs in the interview and discussed in the broader guidance team during reflection meetings.

### Procedure

Potential cases will be recruited through designated contact persons at the four participating institutions and through the adolescents’ individual DBT-A therapists. Based on the eligibility criteria, these contacts will screen all adolescents admitted to DBT-A treatment between 2025 and 2026.

When eligible, the adolescent, their parent(s) and the individual therapist will receive a detailed study information outlining the purpose, procedures, confidentiality measures, voluntary nature of participation and the right to withdraw at any point without consequences. All participants (adolescent, parent(s) and individual therapist) will be asked to sign an informed consent form. A decision period of at least one week will be provided to allow participant sufficient time to consider their involvement. Adolescents under the age of 16 years old will require co-signature by a parent or legal guardian.

Prior to each interview, participants will again be informed about the study procedures, their rights, potential benefits and risks, and the voluntary nature of participation. At each interview session, verbal consent will be reaffirmed. Each semi-structured interview will last approximately 60 minutes and will be conducted either online or on-site at the participating institution, depending on the preference of the participant. Prior to each interview with the adolescent, a brief questionnaire (the Level of Personality Functioning Scale – Brief Form 2.0 (LPFS-BF 2.0) [39] will be completed to assess changes in personality functioning during treatment, as supplement to the qualitative data.

Interviews will be conducted by the lead researcher (AW), under supervision of project team members, who bring extensive expertise in qualitative research (LN) and clinical experience with DBT-A as a child and adolescent psychiatrist (AK). Interviews will be audio-recorded using an Olympus VN-series voice recorder. Verbatim transcripts will be prepared by the lead researcher (AW) and research interns using Microsoft Word. Participants will have the opportunity to review their transcripts and may request (partial) deletion or amendments to statements if desired. Follow up interviews will be scheduled at the conclusions of the first, second, and third interview to ensure continuity. As a token of appreciation, participants (adolescents and parents) will both receive a €50,-gift card upon completion of the final interview. Institutions are financially compensated for the participation of their therapists in the study.

## Materials

### Interview guide

Participants will be asked to reflect on dynamic factors encountered during DBT-A, specifically addressing what they perceived as facilitators and barriers during treatment. The overall aim is to explore how these factors influence the therapeutic process, shape the treatment experience, and contribute to the perceived treatment outcomes, either positively or negatively. Moreover, the role of parents in DBT-A will be explored, focussing on what parents need in order to contribute meaningfully to their child’s DBT-A treatment, how they can (re)establish connection with their child during treatment, and how their involvement relates to the dynamic factors.

The interviews’ topic guide was informed by a pilot study using a stepwise, iterative approach:

1. Literature search to identify theoretically relevant dynamic factors (see appendix S1).
2. Consultation with clinical experts (N=6), all of whom are DBT-A trained therapists working in child and adolescent mental health institutions (Levvel and LUMC Curium). Experts were asked to confirm, clarify, supplement of challenge the findings from the literature search.
3. Consultation with individuals with lived experience (N=4), including three adolescents who had previously completed DBT-A treatment and one parent. These participants were similarly asked to reflect on the relevance and accuracy of the proposed factors.

Based on this process, a semi-structured interview guide was developed deductively, organized around the following themes:

- Motivation for starting DBT-A treatment
- Perceived impact of previous treatment experiences
- Current functioning of the adolescent and family
- Attitudes and responses toward the treatment an treatment process
- Perceptions and experiences related to dynamic factors, including:
  - Therapeutic alliance
  - Ruptures and repair in the therapeutic relationship
  - Communication style
  - Treatment expectations
  - Sense of connection (to self and others)
  - Attitudes of involved parties
  - Emotional responses toward one another
  - Motivation and engagement of the adolescent, parents and therapist
  - Mutual trust and belief in the treatment process
  - Self-disclosure
- Role of parents of DBT-A

During the interviews, probing questions (e.g., “Could you tell me more about…”, “What do you mean by that?” and “How did that make you feel?”). The topic list will be subject to ongoing refinement based on insights gathered from earlier interview rounds, allowing for the inductive inclusion of new themes as they emerge. This iterative approach ensures that the interview guide remains responsive to participants’ lived experiences and our evolving understanding of dynamic factors [40].

### Questionnaires

While the primary focus of this study is on collecting in depth-qualitative data, additional quantitative measures will be used to provide contextual information that supports interpretation of the qualitative findings [41]. This secondary data collection enhances our understanding of each case and contributes to a more comprehensive description of the participant group and to enhance transformability [38]. Relevant clinical and demographic information will be gathered with a short questionnaire during the first interview, including age and gender of the adolescent, family composition, primary DSM-V diagnosis and any comorbid conditions, previous mental health care received. During the individual therapist interview, the following background variables of the therapist will be collected: age and gender, educational background, years of professional experience in child and adolescent psychiatry.

To evaluate the adolescent’s level of personality functioning, LPFS-BF 2.0 [39] will be used. This self-report instrument, aligned with the DSM-5 Alternative Model of Personality Disorders (AMPD) [10], will be administered at all four data collection points during treatment (0, 3, 6, and 9 months). The LPFS-BF 2.0 scores will provide context to the qualitative findings on experienced treatment success. While the design of this study does not allow for causal inferences, a comparison will provide in-depth insights into the developmental course of personality functioning and how it relates to subjective treatment experiences. Additionally, the LPFS-BF facilitates triangulation of how the adolescents report their personal functioning with the perspectives of the parents and individual therapists, contributing to a more holistic understanding of each treatment trajectory.

## Data analysis

A core objective of this study is to examine change over time in the experiences of adolescents, parent(s) and individual therapist during DBT-A treatment. We anticipate that dynamic factors such as the therapeutic relationship, treatment motivation and commitment; and parental involvement will shift throughout the course of therapy. Therefore, in addition to reflexive thematic analysis, we will apply a longitudinal analytic approach that captures both the process and direction of these changes. Our analysis will proceed in three main phases, following principles outlined by Audulv et al. [34] and Grossoehme and Lipstein [42], and guided by Reflexive Thematic Analysis (RTA) methodology [43]. We will analyse within- and across-case, move forward and backward between data from different time points, pose analytical questions regarding time and change and we will use matrices to display change as described by Audulv et al. [34].

### Step 1. Reflexive Thematic Analysis of the interviews

First, we will use RTA to identify and interpret patterns of meaning across narratives. This method enables both inductive (data-driven) and deductive (theory-informed) theme development. Initially, line-by-line coding will be conducted by at least two researchers to ensure inter-coder reliability [44]. Discrepancies will be discussed and resolved collaboratively. Moreover, an expert by experience will read the transcripts and will reflect with AW on the transcripts and codes. At later stages, the primary researcher (AW) will independently code all interviews, with a subset reviewed by supervisors (AK and LN) to ensure consistency. Codes will be grouped into themes using ATLAS.ti V24 [45]. Once key themes are identified, they will be refined into discrete conceptual categories, which will then be analyzed for salience, interrelation, and recurrence across time points. The analysis will be conducted in accordance with the steps described in the methodology of Grossoehme and Lipstein [42]. The analysis in each step will follow the reflective and recursive stages of Byrne [43].

### Step 2. Within-Case matrix

After coding, the data will be organized into individual matrices per trio (adolescent, parent(s), and therapist). In each matrix, themes will be displayed along the Y-axis, and time points (0, 3, 6, and 9 months) along the X-axis (see Table 1: Example of Within-Case matrix). Each cell will include relevant data excerpts from all three participants. This format facilitates comparison within each trio and captures how shared and individual experiences evolve over time.

**Table 1:** Example of a Within-Case matrix.

| Themes | 0 months | 3 months | 6 months | 9 months |
| --- | --- | --- | --- | --- |
| Feeling **motivated** for treatment | Adolescent feels motivated for treatment<br>Parent does not feel motivated for treatment due to a lack of trust in possible treatment success.<br>Therapist feels motivated to treat the adolescent but has doubts on parental involvement. | Adolescent describes a lack of motivation due to a lack of perceived improvement.<br>Parent feels motivated, because of the small positive changes in adolescents behaviour.<br>Therapist still feels motivated to treat the adolescent. | Adolescent feels motivated for treatment because the small improvements are noticeable.<br>Parent feels motivated because of changes in behaviour.<br>Therapist still feels motivated to treat the adolescent. | Participants reflect on the motivation, and all relate the fluctuating motivation to perceived treatment success. |
| Theme B |  |  |  |  |
| Theme C |  |  |  |  |

### Step 3. Cross-Case matrix

A second matrix will then be created to compare experiences across trios. In this matrix, themes— now incorporating their temporal development based on step 2—will form the Y-axis, while each column on the X-axis will represent a specific trio (see Table 2: Example of Cross-Case matrix). Each cell will contain a summary of how the theme developed over time in that case, allowing identification of patterns across participants. Themes may be revised or regrouped into time-sensitive conceptual categories, depending on how they manifest and evolve across different cases.

**Table 2:** Example Cross-Case matrix.

| Themes | Trio 1 | Trio 2 | Trio 3 |
| --- | --- | --- | --- |
| Feeling **motivated** for treatment | No change in feeling motivated for treatment during the course of treatment. | Change from being less motivated to more motivation for the adolescent and parent. Therapist did not change in feeling motivated to treat the adolescent. | Adolescent's motivation fluctuated during therapy<br>Parents motivation positively developed since start treatment.<br>Therapist's motivation did not change. |
| Theme B |  |  |  |
| Theme C |  |  |  |

#### Reflexivity and positionality

Throughout the analytic process, findings and methodological decisions will be systematically discussed during regular team meetings to enhance the rigor and credibility of the study. Moreover, each transcript is discussed based on the reflections of an expert by experience. To ensure transparency and traceability, AW will maintain a detailed research diary documenting analytic decisions, researcher reflections, and team discussions. During regular team meetings, the researchers will engage in systematic reflection on personal, interpersonal, methodological, and contextual reflexivity, as recommended by Olmos-Vega et al. [46].

The research team is composed of professionals with complementary expertise in child and adolescent psychiatry and psychology. AW, a female PhD candidate and psychologist trained in Dialectical Behavior Therapy (DBT), contributes clinical experience as an individual therapist. LN, a female associate professor, offers extensive expertise in qualitative research methodologies. AK, a female psychiatrist, brings a combination of clinical and research experience in child and adolescent psychiatry. This multidisciplinary composition informs both the study’s design and the interpretation of findings, while necessitating ongoing reflexivity to critically examine how the researchers’ professional backgrounds and gender may shape perspectives and analytic processes (personal reflexivity).

The research team acknowledges the importance of therapeutic interventions and the value of qualitative research, and recognizes reflexivity as a fundamental component of the research process. Core values guiding the team include collaboration and equality. Importantly, none of the researchers have direct involvement with the study participants; specifically, AK is not involved with participants at her affiliated institution, thereby upholding interpersonal reflexivity.

The team holds the belief that this research will contribute to the advancement of care practices. Methodological decisions will be made collaboratively, with ongoing reflection on the implications for research quality and ethical standards (methodological reflexivity). It is acknowledged that participation in interviews may prompt participants to reflect on their own treatment, which could positively influence their therapeutic process (contextual reflexivity).

## Ethical considerations

The study protocol has received ethical approval from the Medical Research Ethics Committee Leiden Den Haag Delft (N24.082) prior to the start of data collection. The study will adhere to the ethical principles outlined in the Declaration of Helsinki [47]. To ensure transparency and methodological rigor, the study will adhere to the Consolidated Criteria for Reporting Qualitative Research (COREQ) [48].

### Confidentiality and Data Protection

All data will be pseudonymized to protect the identities of participants. Strict confidentiality will be maintained throughout the research process. The research data will be stored in the secure facilities of LUMC Curium, in accordance with current data security and confidentiality standards.

Importantly, members of the research team will not analyze transcripts from cases associated with the DBT-A teams they are affiliated with, in order to minimize bias and safeguard privacy.

### Safety and Safeguarding Procedures

Should acute and severe concerns regarding a participant’s safety arise during the study, these will be addressed in consultation with an independent researcher and an independent psychiatrist, both of whom are involved in the research project, but not members of any DBT-A treatment team. If necessary—and only after appropriate communication with the adolescent—contact may be made with the individual therapist, the institutional contact person and/or for the adolescent important adult to ensure the participant’s wellbeing. Such actions can potentially impact the participant’s trust and openness toward the researcher in subsequent interviews. In such cases, the research team will formally reflect on and document the incident. Additionally, the adolescent will be invited to engage in a dialogue about issues of openness and trust to promote transparency and to discuss the willingness for further participation.

### Adverse Events

Any adverse or serious events encountered during the research process will be discussed immediately with those involved, reported to the responsible individual therapist or treatment coordinator, logged and monitored by the research team, and reviewed with the head (or sub-department head) of the participating institution, in accordance with the procedures of LUMC Curium.

### Participant Burden and Risk Assessment

Participation involves a modest time investment for interviews and questionnaires. The risks associated with participation are assessed to be minimal, and care has been taken to design the study in a manner that prioritizes participant safety, autonomy, and wellbeing.

## Study timeline

The study spans a total duration of four years, structured across three phases:

### Phase 1: Preparatory activities (8 months)

This initial phase (concluded) focused on laying the groundwork for successful study implementation. Key activities included recruiting and onboarding the PhD candidate (executing researcher); preparing participating clinical partners and informing involved individual therapists; securing all required approvals, including a non-WMO declaration (non-Medical Research Involving Human Subjects Act) from the Medical Ethics Committee of the Leiden University Medical Center (METC-LDD); and approval from the internal Science Committee of LUMC Curium.

### Phase 2: Data collection and analysis (30 months)

The core research phase involves the collection and analysis of longitudinal data. From December 2025 until December 2026 participants will be recruited; semi-structured interviews and questionnaire data will be gathered; preliminary results are expected August 2027 and insights will be discussed with the Youth and Parent Advisory Council of LUMC Curium to ensure that interpretations align with lived experiences; and where possible, intermediate findings will be shared through lectures and internal dissemination at the participating institutions. Moreover, scientific articles will be drafted, and a knowledge dissemination strategy for the final phase will be developed.

### Phase 3: Final analysis and dissemination (10 months)

In the final phase a comprehensive analysis of the full dataset will be conducted; results will be disseminated through academic publications, presentations, and institutional feedback loops; and knowledge sharing will be tailored to both professional and public audiences.

## Discussion

Synthesis of the results of this longitudinal, multi-perspective analyses will offer nuanced insight into how dynamic factors unfold over time in DBT-A and contribute to perceived treatment success.

These findings will inform both clinical practice and future research on mechanisms underlying perceived treatment success in DBT-A. Alongside two to three scientific papers in peer reviewed journals, dissemination will take place by presenting at national and international conferences, Dutch-language handouts including key takeaways for clinical practice, and hosting lectures at the participating institutions. Where feasible, relevant information will be shared at institutions where DBT-A training is given. Also, key findings will be translated in collaboration with the Dutch National Youth Council (NJR), to promote knowledge transfer towards adolescents and their parents.

### Limitations and challenges

While the study design is robust in its longitudinal, multi-perspective approach, several practical limitations must be acknowledged. First, adolescents in DBT-A are often in crisis or emotionally overwhelmed at treatment onset. Their mental health challenges, combined with the intensity of DBT-A, may reduce their capacity or willingness to participate in research. To address this and minimize participant burden, participants will be recruited by their therapists, and interviews will be flexibly scheduled – including online options and times outside of school or therapy hours. Second, parents may be experiencing significant stress and emotional fatigue due to the severity of their child’s condition. Efforts will be made to support parents’ engagement, emphasizing the potential contribution their perspectives make to the improvement of care. Third, involving individual therapists presents its own set of challenges. High caseloads and time constraints may affect their willingness to participate. Additionally, some therapists may find it uncomfortable to reflect on their client’s experience in a research setting. To mitigate these challenges, the research team will stress to all participants that participation is voluntary and that interviews are confidential: information shared by one participant will not be disclosed to the others.

Another challenge known with a longitudinal design is incomplete follow-up of participants and attrition with loss to follow-up over time [49]. If a participant discontinues treatment but still gives consent for the research project, the data is still included, as it is considered as valuable information on potential treatment failure.

Any amendments that occur during the research project will be discussed with the research team and with the contact persons of the participating institutions. The changes for early termination of the study are considered low, therefore there is no termination plan for early termination.

## Conclusion

This qualitative, longitudinal, and multi-perspective study aims to provide an in-depth understanding of how dynamic factors within DBT-A develop and influence treatment outcomes for adolescents with BPD features/traits. By integrating the perspectives of adolescents, their parents, and individual therapists across multiple time points, the study will offer novel insights into the development of dynamic factors and underlying therapeutic change in DBT-A. These findings will contribute to tailored and responsive interventions, inform clinical practice, and support the development of training and policy for DBT-A. Ultimately, this study seeks to improve treatment outcomes and engagement in DBT-A for adolescents and their families.

## Data Availability

All relevant data are within the manuscript and its Supporting Information files.

## Authors’ contributions

*Conceptualization*: Laura Nooteboom, Anne Krabbendam, Agaath Koudstaal, Joep Sins, Jacquelijne Schraven, Robert Vermeiren, Jantine Roeleveld

*Methodology*: Laura Nooteboom and Anne Krabbendam

*Funding acquisition*: Laura Nooteboom, Anne Krabbendam, Agaath Koudstaal, Joep Sins, Jacquelijne Schraven, Robert Vermeiren, Jantine Roeleveld

*Writing-First draft*: Anneke de Weerd and Laura Nooteboom

*Writing-Review and editing*: Anne Krabbendam, Agaath Koudstaal, Joep Sins, Jacquelijne Schraven, Robert Vermeiren, Jantine Roeleveld, Elisabeth Koopman-Verhoeff, Laura Nooteboom

## Acknowledgements

The authors would like to thank the involved clinical experts, adolescents and parents with experience for participating in the pilot study to design the topic list. The authors would also like to thank all members of the DBT-A team of the participating institutions for willing to participate.

This study is funded by the Dutch ‘Stichting tot Steun VCVGZ’ under project code p313.

## Supporting Information

S1. Reference list topic list (pilotstudy)

